# Immunoglobulin G glycome composition in transition from pre-menopause to menopause

**DOI:** 10.1101/2021.04.10.21255252

**Authors:** Domagoj Kifer, Helena Deriš, Ana Cindrić, Tea Petrović, Ana Cvetko, Irena Trbojević-Akmačić, Louise Newson, Tim Spector, Cristina Menni, Gordan Lauc

## Abstract

**Background:** Glycosylation of immunoglobulin G (IgG) is an important regulator of the immune system and its changes are believed to be a significant contributor to inflammaging. Gonadal hormones affect IgG glycome composition, suggesting that alterations in IgG glycosylation might be one of the molecular mechanisms behind increased disease risk in perimenopause.

**Methods:** IgG was isolated from 5,354 plasma samples collected from 1,940 females and 113 males at multiple time points. IgG glycans were released, labelled with a fluorescent dye and analysed by ultra-high-performance liquid chromatography. Mixed modelling was used to determine average levels of individual IgG glycans in pre-menopausal women, menopausal women, and men.

**Findings:** Large and statistically significant differences in IgG glycome composition were observed, mainly reflecting decreased galactosylation and sialylation of glycans in menopausal women. During perimenopause women had a significant higher rate of increase in agalactosylated structures (0.051/yr; 95%CI = 0.043 - 0.059, p<0.001), and decrease in digalactosylated (−0.043/yr; 95%CI = -0.050 – -0.037, p<0.001), and monosialylated glycans (- 0.029/yr; 95%CI = -0.034 – -0.024, p<0.001), compared to premenopausal women.

**Interpretation:** Proinflammatory IgG glycome and the resulting decrease in the ability of IgG to suppress low-grade chronic inflammation may be an important molecular mechanism mediating the increased health risk in perimenopause. IgG glycome changes considerably during perimenopause and may aid the diagnosis of perimenopause.

**Funding:** Croatian National Centre of Excellence in Personalised Healthcare, ESI Funds grant for the Centre of Competences in Molecular Diagnostics, the Wellcome Trust and the Medical Research Council (MRC)/British Heart Foundation (BHF).

## Introduction

Glycosylation of immunoglobulin G (IgG) is an important regulator of the immune system ^1^. In addition to directly affecting the effector functions of IgG by promoting its binding to different Fc receptors ^2^, glycans attached to IgG have numerous other roles in the regulation of the immune system ^3,4^. Glycans are also important mediators of the anti-inflammatory activity of IgG ^5,6^ as well as the driving force for one of the molecular mechanisms underlying immunosuppressive activity of intravenous IgG (IVIG) therapy used to treat a variety of immunological disorders ^7^. Changes in IgG glycosylation are also believed to be an important contributor to aging at the molecular level through a process called inflammaging ^8^.

IgG glycome composition is tightly regulated and, despite the absence of a direct genetic template, is heritable to a significant extent ^9,10^. Alternative glycosylation (the attachment of different glycan structures to the same glycosylation site) is structurally and functionally analogous to coding mutations, but instead of being defined by sequence variation in individual genes, glycome composition is inherited as a complex trait encoded by multiple genes ^11^. Genome-wide association studies identified a network of over 30 genes that associate with the IgG glycome composition ^12–14^. One of the regulators of IgG glycosylation are sex hormones, and the association between estradiol and the IgG glycome composition was confirmed to be causal ^15^. Recent randomised placebo-controlled clinical study demonstrated that the deprivation of gonadal hormones resulted in an increase of biological age measured by IgG glycans, which was completely prevented by estradiol supplementation ^16^.

Large population studies revealed that IgG glycome composition changes with age and that in females this change is particularly pronounced in the time preceding the average age of menopause ^17,18^. However, the association between changes in the IgG glycome composition and menopause has not yet been validated. Perimenopause is accompanied by different symptoms and is very often misdiagnosed due to changing hormone levels and symptom overlap with other conditions. Menopause associates with an increased risk of developing a range of diseases affecting metabolism, bones, cardiovascular system, brain, even cancer ^19^. Early perimenopause diagnosis would enable timely interventions to ease the symptoms, decrease risks of developing accompanying diseases, and timely monitoring of any changes in women’s health status due to hormonal changes. To fill this knowledge gap and explore the association of IgG glycome changes with perimenopause, we analysed IgG N-glycome composition in 5,080 samples from 1,940 females in multiple time points during their transition from pre-menopause to menopause, along with 274 samples from 113 males as an additional control.

## Methods

### Study population

Study subjects were individuals enrolled in the TwinsUK registry, a national register of adult twins recruited as volunteers without selecting for any particular disease or traits ^20^. Data on menopause was gathered through the multiple choice questionare asking women at each visit: “What is your menopausal status?” with possible answers “Pre menopausal”, “Going through the menopause”, “Post menopausal” and “Don’t know” and “How old were you when you became post-menopausal (when you stopped having periods for one year or more)?”. Consequently sample was defined as menopausal if questionar resulted with “Post-menopausal” answer or age larger then reported age of becoming postmenopausal. Similarly sample was defined as premenopausal if questionare resulted with “Pre menopausa” answer of age at sampleing was less then reported age of becoming postmenopausal reduced by 1 year. Any inconsistent (i.e. “Pre menopausal” answer at age after becoming menopausal) or unreliable data (i.e. answered as “Going through the menopause”) were dropped from the study (Supplementary Table 1). The TwinsUK study was approved by NRES Committee London–Westminster, and all twins provided informed written consent.

### Isolation of IgG

IgG isolation from serum samples was performed on protein G monolithic 96-well plates as previously described ^21^. In brief, 100 µL of serum was diluted 1:7 (v/v) with PBS (phosphate buffer saline) followed by filtration on a 0.45 µm GHP filter plate (Pall Corporation, USA). Diluted and filtered serum samples were then transferred to the protein G monolithic plate, which was then washed. Elution of IgG was performed with 1 mL of 0.1 mol/L formic acid (Merck, Germany) followed by immediate neutralization of the mixture with ammonium bicarbonate (Acros Organics, USA) to pH 7.0. IgG-containing eluate was aliquoted into PCR plate (average mass of 15 µg of IgG per sample) and dried in a vacuum centrifuge.

### Deglycosylation, labelling and purification of IgG N-glycans

Deglycosylation, RapiFluor-MS labelling and purification of IgG N-glycans were performed using the GlycoWorks RapiFluor-MS N-Glycan Kit obtained from Waters Corporation (USA). The entire procedure was done following Waters’ protocol (Waters Corporation 2017, https://www.waters.com/webassets/cms/support/docs/715004793en.pdf). In the end, the samples of released and labelled IgG glycans were stored at -20 °C until further use.

### HILIC-UHPLC-FLR analysis of RapiFlour-MS labelled IgG N-glycans

RapiFluor-MS labelled IgG N-glycans were analysed using ultra-high-performance liquid chromatography based on hydrophilic interactions with fluorescence detection (HILIC- UHPLC-FLR) on Waters Acquity UPLC H-class instruments. The instruments were controlled and monitored with the Empower 3 software, build 3471 (Waters, USA). Chromatographic separation of glycan structures was performed on Waters UPLC Glycan bridged ethylene hybrid (BEH) Amide chromatographic columns (130 Å, 1.7 µm BEH particles, 2.1×100 mm). Solvent A was 50 mmol/L ammonium formate, pH 4.4, while solvent B was 100% LC-MS grade acetonitrile. A linear gradient of 75–61.5 % acetonitrile (v/v) was used at a flow rate of 0.4 mL/min over 30 minutes in a 42-minute analytical run. Each chromatogram was separated into 22 glycan peaks containing IgG glycan structures (**Supplementary Figure 1**.), by automated integration ^22^ resulting in relative quantification of IgG N-glycans (total area normalization). Specific N-glycan structures found in each glycan peak according to Keser et al. ^23^ are presented in **Supplementary Table 2**. Formulas used for the calculation of IgG derived traits are presented in **Supplementary Table 3**.

### Statistical analysis

After UHPLC analysis, each measured glycan peak was normalized by total chromatogram area. Observed relative glycan abundances were log transformed and batch corrected using ComBat method (R package ‘sva’ ^24^). After batch correction, values were back-transformed to relative abundance and derived traits were calculated according to **Supplementary Table 3**.

Overlapping measured N-glycomes and information available on menopause found in TwinsUK register, resulted in 5354 samples from 1940 females and 113 males.

For each glycan trait the effect of menopause on the abundance was estimated by mixed modelling (R package ‘lme4’ ^25^). Logit transformed relative abundance was used as dependent variable and sex, menopausal status (nested within sex), age and age- menopausal status interaction as fixed factors, as well as family ID and individual ID (nested within family ID) as random intercepts and age as random slope. Using model, mean values were estimated for each group and pairwise compared using t-test (post-hoc) (R package ‘emmeans’ ^26^). Given multiple testing for large number of glycans, the control of false discovery rate (FDR) was done by adjusting p values according to Benjamini-Hochberg’s method.

For the analysis of the rate of change of glycan abundances, average yearly changes were calculated from two timepoints according to the formula: rate = (logit(glycan_2_) – logit(glycan_1_))/(age_2_ – age_1_), where glycan is relative abundance resulted from normalized chromatogram, and age is represented in years, assuming age_2_ > age_1_. Depending on menopausal status in the first and second timepoint measurements were classified into three categories: premenopausal (both timepoints before menopause), perimenopausal (first timepoint before, and second in the menopause), and menopausal (both timepoints in menopause). Mean rates were estimated by the mixed model, in which rate was dependent variable, and sex, menopausal group (nested within menopausal status) and mean age (calculated as (age_2_+age_1_)/2) were set as fixed factors, as well as family ID and individual ID (nested within family ID) as random intercepts. Estimated mean rates for each menopausal group and males were pairwise compared using t-test (post-hoc). FDR control was done by adjusting p values according to Benjamini-Hochberg’s method.

N-glycome (observed in one timepoint) potential for diagnosis of perimenopause was analysed in a subset of women aged between 45 and 55 years by logistic regression with L1 regularization technique (R package ‘caret’ and ‘glmnet’ ^27,28^). The subset was split into training (randomly chosen one measurement per family) and test (everything else) subset. The logistic model was defined with menopausal status (TRUE/FALSE) as the dependent variable, and age and all directly measured glycans as independent variables. To avoid overfitting, coefficients of the logistic model were estimated by 10-fold cross validation on training subset data, with hyperparameter lambda set to lambda_min_. The model was tested on test subset; results are presented by receiver operating characteristic (ROC) curve. In parallel, the null model was fitted in the same way but using only age as a predictor. Observed ROC curves of the two models were compared by a bootstrap test (B = 2000) (R package ‘pROC’ ^29^).

The rate of N-glycome change (based on two timepoints) potential for diagnosis of perimenopause was analysed in a subset of women with median two-timepoint age between 45 and 55 years in the same way as diagnosis of menopause with IgG glycome analysed in one timepoint. The perimenopausal category was defined as TRUE, while others (menopausal and premenopausal) were defined as FALSE. All statistical analyses were done using R ^30^.

## Results

IgG glycome composition was analysed in multiple samples from 1,940 females and 113 males from the UK registry of adult twins. The 1,087 samples from 732 individual twins (500 families) were collected prior to the onset of menopause, while 3,993 samples from 1,678 individual twins (969 families) were collected after the onset of menopause (Supplementary Table 1). Additionally, IgG glycome was analysed in 113 men (274 samples) as an additional control. A representative chromatogram with detailed structures of individual IgG glycans is shown in Supplementary Figure 1. Mixed modelling was used to determine average levels of individual IgG glycans in pre-menopausal women, menopausal women, and men. Large and statistically significant differences were observed in multiple glycans, mainly reflecting decreased galactosylation and sialylation of glycans in menopausal women (Figure 1). Levels of individual IgG glycans are available in Supplementary Figure 2.

**Figure 1.**
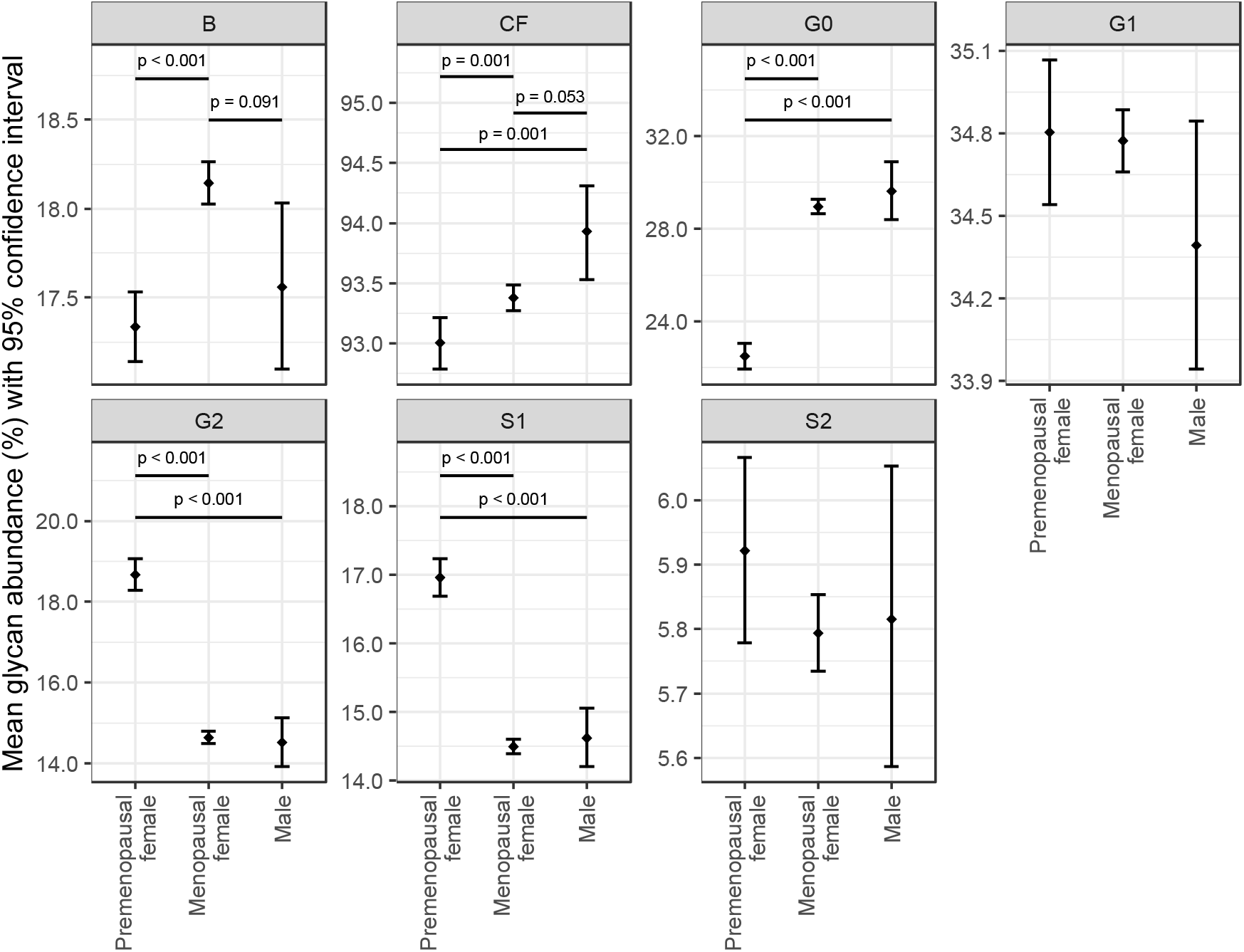
IgG glycome composition in premenopausal women, menopausal women, and men. Mean glycan abundances (% of total IgG glycome) and corresponding 95% confidence intervals were estimated from fitted mixed model with logit transformed glycan as dependent variable and sex, menopausal status (nested within sex), age and age-menopausal status interaction as fixed factors, as well as family ID and individual ID (nested within family ID) as random intercepts and age as random slope. Only adjusted post-hoc p values less than 0.1 are shown. B - bisecting N-acetylglucosamine (GlcNAc), CF - core fucose, G0 - agalactosylated glycans, G1 - monogalactosylated glycans, G2 - digalactosylated glycans, S1 - monosialylated glycans, and S2 - disialylated glycans.

It is known that IgG galactosylation and sialylation decrease with age ^17^ thus we attempted to evaluate whether the changes in glycans associated with the transition to menopause were more extensive than the age-related changes. Since we had multiple samples from the same individual, we were able to calculate the rate of change of individual IgG glycans through time. A subset of women in our cohort (n = 379) entered menopause between two sampling time points, thus for them, we were also able to calculate the rate of changes in individual IgG glycans during the perimenopausal period. The comparison of the rate of age-related IgG glycome changes (when there was no change of the menopause status) with the rate of IgG glycome changes during perimenopause (i.e., between time points when the transition to menopause occurred) revealed statistically significant differences for a number of glycans (Supplementary Figure 3). The most prominent differences in the perimenopause period were the significantly larger rates of increase in the agalactosylated structures (G0) and the decrease of digalactosylated (G2) and monosialylated (S1) glycans (Figure 2). During perimenopause women had a significant higher rate of increase in agalactosylated structures (0.051/yr; 95%CI = 0.043 - 0.059, p<0.001), and decrease in digalactosylated (- 0.043/yr; 95%CI = -0.050 – -0.037, p<0.001), and monosialylated glycans (−0.029/yr; 95%CI = -0.034 – -0.024, p<0.001), compared to premenopausal women.

**Figure 2.**
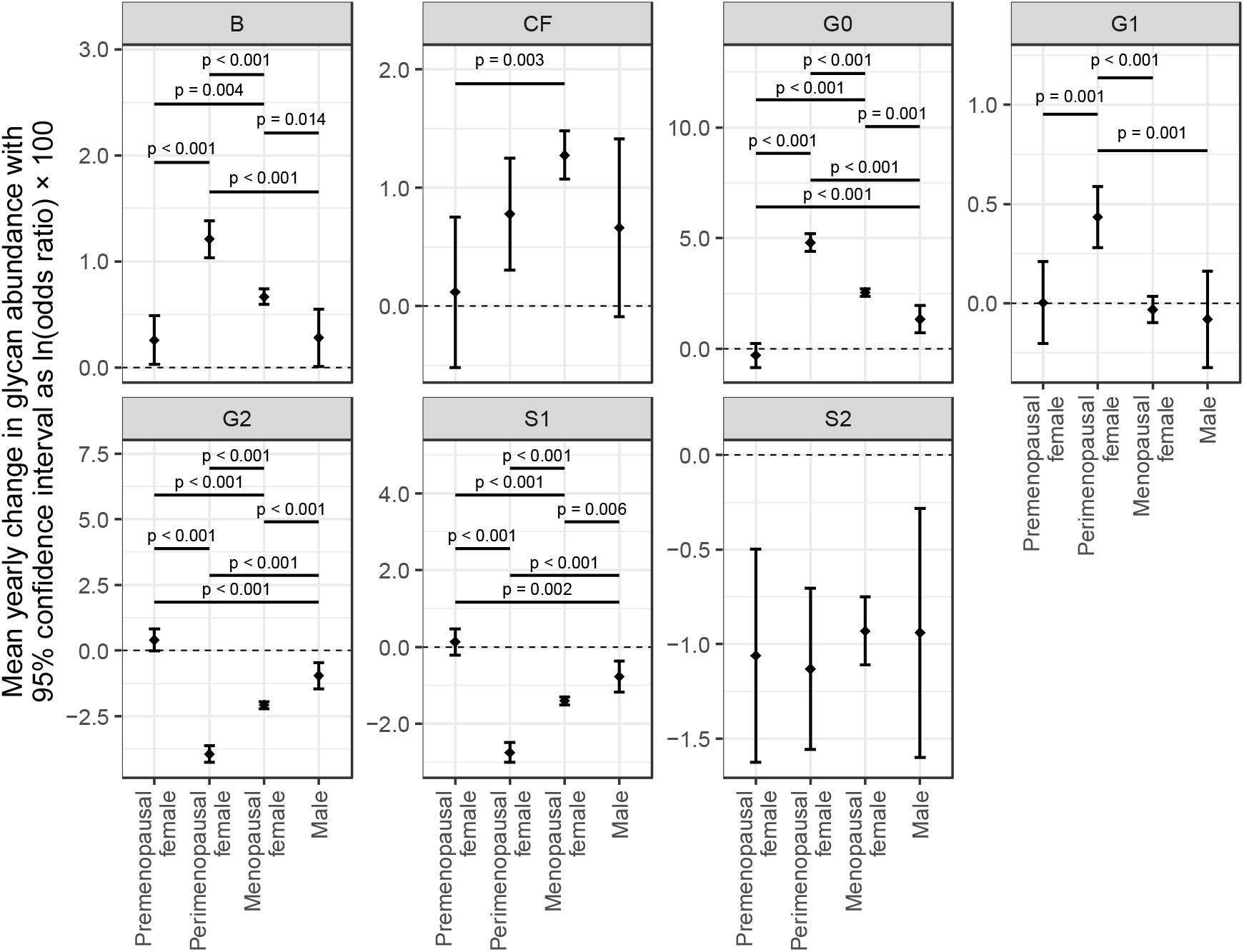
Average yearly change of IgG glycans in females during the perimenopause period, females in pre- or post-menopause, and males. Mean yearly change in IgG glycan abundances and corresponding 95% confidence intervals were estimated from fitted mixed model with logit transformed glycan as dependent variable and sex, menopausal status (nested within sex), age and age-menopausal status interaction as fixed factors, as well as family ID and individual ID (nested within family ID) as random intercepts and age as random slope. Only adjusted post-hoc p values less then 0.1 are shown. B - bisecting GlcNAc, CF - core fucose, G0 - agalactosylated glycans, G1 - monogalactosylated glycans, G2 - digalactosylated glycans, S1 - monosialylated glycans, and S2 - disialylated glycans.

Using single-point measurements of IgG N-glycans we attempted to predict menopause status. The lasso model was fitted with menopausal status (TRUE for those who entered menopause and FALSE for all others) as dependent variable and age and all directly measured IgG glycans as predictors. The model was fitted on a randomly chosen one twin per family (training set) in the subset of women with a two-timepoint median age between 45 and 55 years. The remaining twins were used for testing the fitted model. The same train and test subset model with only mean age as a predictor were used to fit and test the model for comparison (Figure 3A). IgG N-glycome in combination with age measured in a single timepoint has shown a good classification performance, with the AUC of 0.853 (95% CI 0.823-0.880) outperforming only age as a predictor of perimenopause (AUC of 0.818, 95% CI 0.783-0.849).

**Figure 3..**
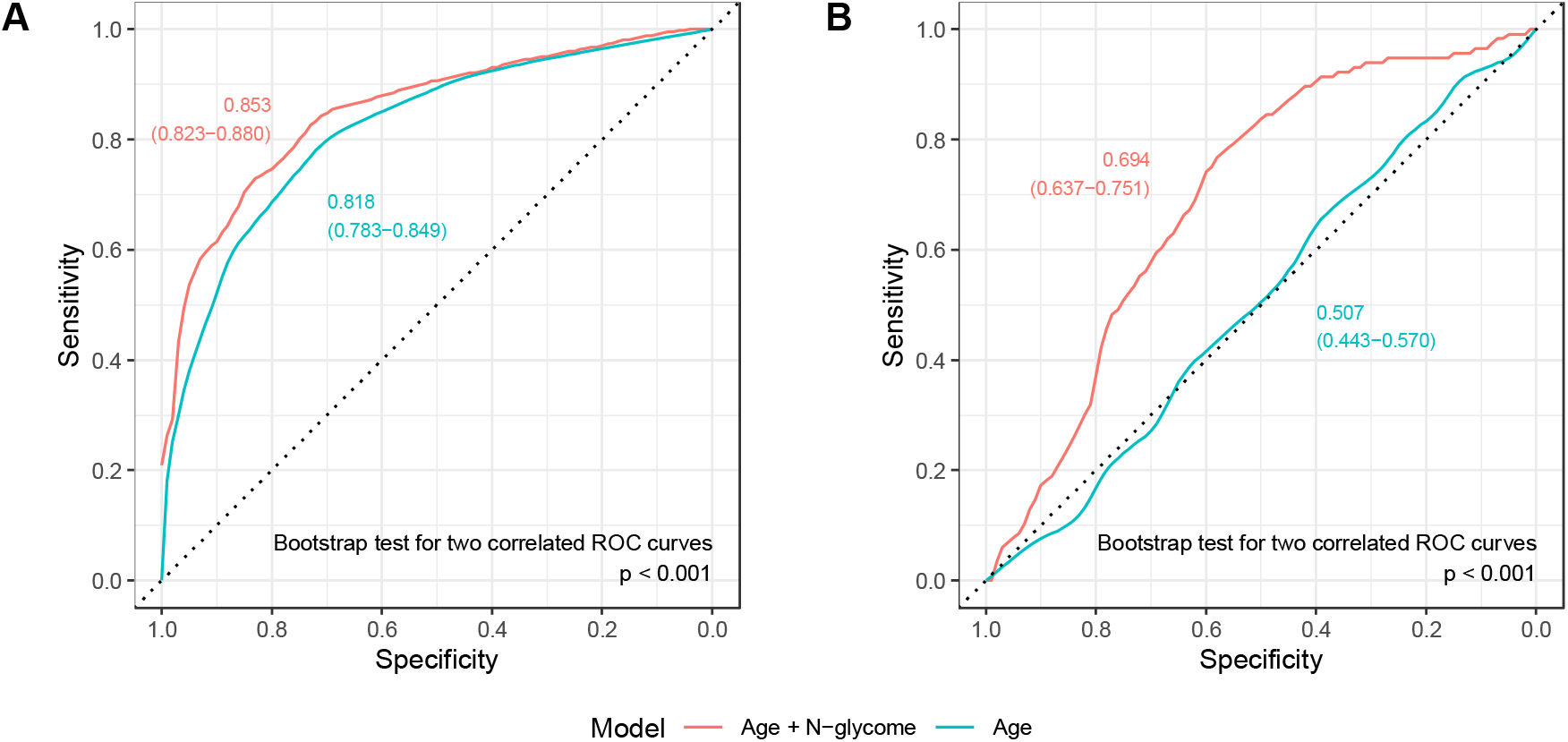
Receiver operating characteristic (ROC) curves for the prediction of perimenopause using IgG glycans measured in a single time point (A) and the change in IgG glycan levels between the two timepoints (B). Graphs are showing area under the curve (AUC) values with 95% confidence intervals.

Since changes in the IgG glycome composition were the most prominent in te period of perimenopause, we used the rate of changes in glycans to predict whether a given women already entered the period of perimenopause, or not Figure 3B). The model was fitted on a randomly chosen one twin per family (training set) in the subset of women with a two-timepoint median age between 45 and 55 years. The remaining twins were used for testing the fitted model. The addition of the information about changes in the IgG glycan levels to a prediction model notably increases AUC value compared to using only age (0.694, 95% CI 0.637-0.751 compared to 0.507, 95% CI 0.443-0.570, respectively).

## Discussion

In this study we have shown the association between the period of perimenopause and the extensive changes in the IgG glycome composition. Aging continuously associates with the transition of the IgG glycome from an inflammation-suppressive to a proinflammatory composition, but this change is much faster in the period when women are progressing from a regular cycle to menopause. The depletion of estrogen during perimenopause and menopause is an important event of aging since low hormone levels increase vulnerability to diseases in hormone-responsive tissues, including the brain, bone, and the cardiovascular system. Menopause is therefore more complex than just being a natural condition resulting in symptoms; it needs to be considered as a long-term hormone deficiency leading to health risks if not appropriately managed with lifestyle adjustments or hormone replacement therapy (HRT) ^31^. After entering menopause, there is an increased risk of developing obesity and metabolic disease, cardiovascular diseases, osteoporosis and arthritis, dementia and cognitive decline, and cancer ^19^.

Previous population studies revealed that IgG N-glycome composition changes with age, especially in females in the time preceding the average age of menopause ^17,18^. In this study we used multiple samples from the same individuals to confirm the association between perimenopause and the extensive changes in the IgG glycome composition. The most extensive change in the IgG glycome during perimenopause was a decrease in galactosylation and sialylation, a change that is generally associated with aging ^1^. Aging continuously associates with the transition of the IgG N-glycome from an inflammation-suppressive to a proinflammatory composition, but this change is much faster in the period when women are progressing from a regular cycle to menopause.

The age-related gradual decrease in the level of galactosylated IgG and the build-up of agalactosylated IgG is known to exacerbate low-grade chronic inflammation ^32–34^ and acts as an effector of pro-inflammatory pathological changes ^35,36^. Decreased levels of IgG galactosylation and/or sialylation have been associated with a wide range of autoimmune conditions, such as systemic lupus erythematosus, inflammatory bowel disease, and many others. Changes in these IgG glycosylation traits are also associated with disease progression, disease activity, symptom severity, and response to treatment. In many cases, these changes occur before the onset of symptoms ^37^.

The rate of change from “young” to “old” IgG glycome increased significantly in the perimenopausal period, suggesting that the loss of immunosuppressive functions of IgG may be an underlying molecular mediator of at least some health risks in this period. The results of this study also indicate the potential of the IgG glycome composition as a biomarker for perimenopause. Perimenopause can last up to 15 years and is difficult to diagnose due to highly irregular hormonal cycles ^38^. As a result of poor awareness and inappropriate use of hormonal tests, women are often misdiagnosed with conditions such as fibromyalgia, migraines, depression, or chronic fatigue syndrome and are frequently prescribed antidepressants despite there being no evidence to support their use to improve the low mood associated with perimenopause or menopause ^39^. The importance of early diagnosis of perimenopause is supported by the research which has shown that early use of HRT leads to a greater reduction of future disease incidence as well as minimising the duration of symptoms ^40^.

Recent placebo-controlled randomised trials indicated the importance of estrogens in the regulation of IgG glycosylation ^15,41^, thus it seems quite probable that the decrease in estrogen levels is an important driver of changes in IgG glycosylation in primenopause. Contrary to estrogens that have a very short half-life in circulation, IgG has a half-life of three weeks ^42^, which implies that the current composition of the IgG glycome is affected by average concentrations of estrogen in the past 3-4 weeks. Since changes in the IgG glycome during perimenopause are so extensive, they could potentially be used as a diagnostic tool. In fact, IgG glycome composition could be used to determine long-term average concentrations of sex hormones in the same way glycated hemoglobin (HbA1c) is used to determine long-term concentrations of blood glucose.

### Limitations of the study

The main limitation of this study is relatively long time between individual sampling time points (6.9±2.6 years) that was not tailored for the occurrence of menopause. Therefore, in some cases, perimenopause was just a short part of the period in which the change in the IgG N-glycome occurred, while in others sampling might have occurred within the perimenopause period, with part of the perimenopause being before and another part after a specific sampling. Hopefully, a future study that would have more frequent sampling during the perimenopause period would enable an even more accurate assessment of IgG glycome association with perimenopause.

## Conclusions

Changes in IgG glycosylation from “young” to “old” glycome associate with many health risks that accompany menopause ^1^. In some diseases like rheumatoid arthritis ^43,44^ and cardiovascular diseases ^45,46^ this change was shown to occur years before disease onset, suggesting the causal role of IgG glycans in disease development. Proinflammatory IgG glycome and the resulting decrease in the ability of IgG to suppress low-grade chronic inflammation may be an important molecular mechanism that mediates the increased health risk in perimenopause, thus this topic should be studied in more detail.

## Data Availability

Data is available upon a reasonable request.

## Acknowledgements

This study has been supported by the Croatian National Centre of Excellence in Personalised Healthcare (Contract no. KK.01.1.1.01.0010), ESI Funds grant for the Centre of Competences in Molecular Diagnostics and the Human Glycome Project. Equipment and products from Waters and New England Biolabs were used for this research. The Department of Twin Research receives support from grants from the Wellcome Trust (212904/Z/18/Z) and the Medical Research Council (MRC)/British Heart Foundation (BHF) Ancestry and Biological Informative Markers for Stratification of Hypertension (AIM-HY; MR/M016560/1), European Union, Chronic Disease Research Foundation (CDRF), Zoe Global Ltd., the NIHR Clinical Research Facility and Biomedical Research Centre (based at Guy’s and St Thomas’ NHS Foundation Trust in partnership with King’s College London). C.M. is funded by the Chronic Disease Research Foundation and by the MRC AIM-HY project grant.

## Supplementary Tables and Figures

**Supplementary Table 1.** Descriptive information about the cohort

|  | Samples | Individuals | Families |
| --- | --- | --- | --- |
| Count, N | 5354 | 2053 | 1173 |
| - by sex (%) |  |  |  |
| Male | 274 (5) | 113 (6) | 69 (6) |
| Female | 5080 (95) | 1940 (94) | 1104 (94) |
| - by group (%) |  |  |  |
| Pre-menopause | 1087 (21) | 732 | 500 |
| Menopause | 3993 (79) | 1678 | 969 |
| - by age subset (%) |  |  |  |
| Between 45 and 55 years | 1374 (26) | 1065 (52) | 670 (57) |
| - by sex (%) |  |  |  |
| Male | 87 (6) | 59 (6) | 39 (6) |
| Female | 1287 (94) | 1006 (94) | 631 (94) |
| - by group (%) |  |  |  |
| Pre-menopause | 507 (39) | 453 | 340 |
| Menopause | 780 (61) | 662 | 458 |
| - by number of samples (%) |  |  |  |
| One |  | 203 (10) |  |
| Two |  | 399 (19) |  |
| Three |  | 1451 (71) |  |
| Age in years, mean (SD) | 58.3 (11.3) | 57.6 (10.3) | 57.2 (10.3) |
| - by sex |  |  |  |
| Male | 57.9 (11.7) | 57.4 (10.8) | 57.4 (10.9) |
| Female | 58.3 (11.2) | 57.6 (10.3) | 57.2 (10.3) |
| - by group |  |  |  |
| Pre-menopause | 43.1 (7.1) | 44.0 (6.0) | 44.6 (5.9) |
| Menopause | 62.4 (8.2) | 61.7 (6.6) | 61.3 (6.7) |
| - by age subset |  |  |  |
| Between 45 and 55 years | 50.6 (3.1) | 50.7 (2.6) | 50.6 (2.6) |
| - by sex |  |  |  |
| Male | 50.5 (3.1) | 50.6 (2.6) | 50.6 (2.5) |
| Female | 50.6 (3.1) | 50.7 (2.6) | 50.6 (2.6) |
| - by group |  |  |  |
| Pre-menopause | 48.5 (2.6) | 48.5 (2.4) | 48.7 (2.4) |
| Menopause | 51.9 (2.7) | 52.2 (2.3) | 52.0 (2.2) |
| Follow up time in years, mean (SD)<br>between consecutive samples |  | 6.94 (2.58) |  |
| - by sex |  |  |  |
| Male |  | 7.17 (2.94) |  |
| Female |  | 6.93 (2.56) |  |
| total follow up time |  | 12.2 (4.3) |  |
| - by sex |  |  |  |
| Male |  | 12.1 (4.8) |  |
| Female |  | 12.2 (4.3) |  |

**Supplementary Table 2.** Detailed description of glycan structures according to Keser et al. ^23^ corresponding to every individual immunoglobulin RapiFluor-MS glycan peak.*

| **RapiFluor-MS glycan peak** | **Glycan structure** | **Description** | **Formula** |
| --- | --- | --- | --- |
| GP1 | FA1 | core fucosylated, monoantennary | GP1 / GP |
| GP2 | A2 | agalactosylated, biantennary | GP2 / GP |
| GP3 | FA2;<br>A2B | core fucosylated, biantennary (major);<br>biantennary with bisecting GlcNAc (minor) | GP3 / GP |
| GP4 | FA2B;<br>M5 | core fucosylated, biantennary with bisecting GlcNAc (major);<br>oligomannose (minor) | GP4 / GP |
| GP5 | A2[6]G1 | monogalactosylated, biantennary | GP5 / GP |
| GP6 | A2[3]G1 | monogalactosylated, biantennary | GP6 / GP |
| GP7 | FA2[6]G1 | core fucosylated and monogalactosylated, biantennary | GP7 / GP |
| GP8 | FA2[3]G1 | core fucosylated and monogalactosylated, biantennary | GP8 / GP |
| GP9 | FA2[6]BG1 | core fucosylated and monogalactosylated, biantennary with<br>bisecting GlcNAc | GP9 / GP |
| GP10 | FA2[3]BG1 | core fucosylated and monogalactosylated, biantennary with<br>bisecting GlcNAc | GP10 / GP |
| GP11 | A2G2 | digalactosylated, biantennary | GP11 / GP |
| GP12 | FA2G2;<br>A2BG2 | core fucosylated, digalactosylated, biantennary (major);<br>digalactosylated, biantennary with bisecting GlcNAc (minor) | GP12 / GP |
| GP13 | FA2BG2 | core fucosylated, digalactosylated, biantennary with bisecting<br>GlcNAc | GP13 / GP |
| GP14 | FA2[6]G1S1;<br>FA2[3]G1S1 | core fucosylated, monogalactosylated and monosialylated<br>biantennary;<br>core fucosylated, monogalactosylated and monosialylated<br>biantennary | GP14 / GP |
| GP15 | A2G2S1 | digalactosylated and monosialylated biantennary | GP15 / GP |
| GP16 | FA2G2S1 | core fucosylated, digalactosylated and monosialylated<br>biantennary | GP16 / GP |
| GP17 | FA2BG2S1 | core fucosylated, digalactosylated and monosialylated<br>biantennary with bisecting GlcNAc | GP17 / GP |
| GP18 | n.d. | structure not determined | GP18 / GP |
| GP19 | A2G2S2 | digalactosylated and disialylated biantennary | GP19 / GP |
| GP20 | A2BG2S2 | digalactosylated and disialylated biantennary with bisecting<br>GlcNAc | GP20 / GP |
| GP21 | FA2G2S2 | core fucosylated, digalactosylated and disialylated<br>biantennary | GP21 / GP |
| GP22 | FA2BG2S2 | core fucosylated, digalactosylated and disialylated<br>biantennary with bisecting GlcNAc | GP22 / GP |

\*structure abbreviations – all N-glycans have two core *N*-acetylglucosamines (GlcNAcs); F at the start of the abbreviation indicates a core-fucose $\alpha$ 1,6-linked to the inner GlcNAc; Mx, number (x) of mannose on core GlcNAcs; Ax, number of antenna (GlcNAc) on trimannosyl core; A2, biantennary with both GlcNAcs as $\beta$ 1,2-linked; B, bisecting GlcNAc linked $\beta$ 1,4 to $\beta$ 1,3 mannose; G(x), number (x) of $\beta$ 1,4 linked galactose on antenna; S(x), number (x) of sialic acids linked to galactose

**Supplementary Table 3.** Formulas used for the calculation of IgG derived traits

| ***Structural feature*** | ***Formula*** |
| --- | --- |
| Agalactosylation (G0) | GP1+GP2+GP3+GP4 |
| Monogalactosylation (G1) | GP5+GP6+GP7+GP8+GP9 |
| Digalactosylation (G2) | GP10+GP11+GP12+GP13 |
| Monosialylation (S1) | GP14+GP15+GP16+GP17 |
| Disialylation (S2) | GP19+GP20+GP21+GP22 |
| Incidence of bisecting GlcNAc (B) | GP4+GP9+GP10+GP13+GP17+GP20+GP22 |
| Core fucosylation (CF) | GP1+GP3+GP4+GP7+GP8+GP9+GP10+GP12+GP13+GP14+GP16+GP17+GP21+GP22 |

**Supplementary Figure 1.**
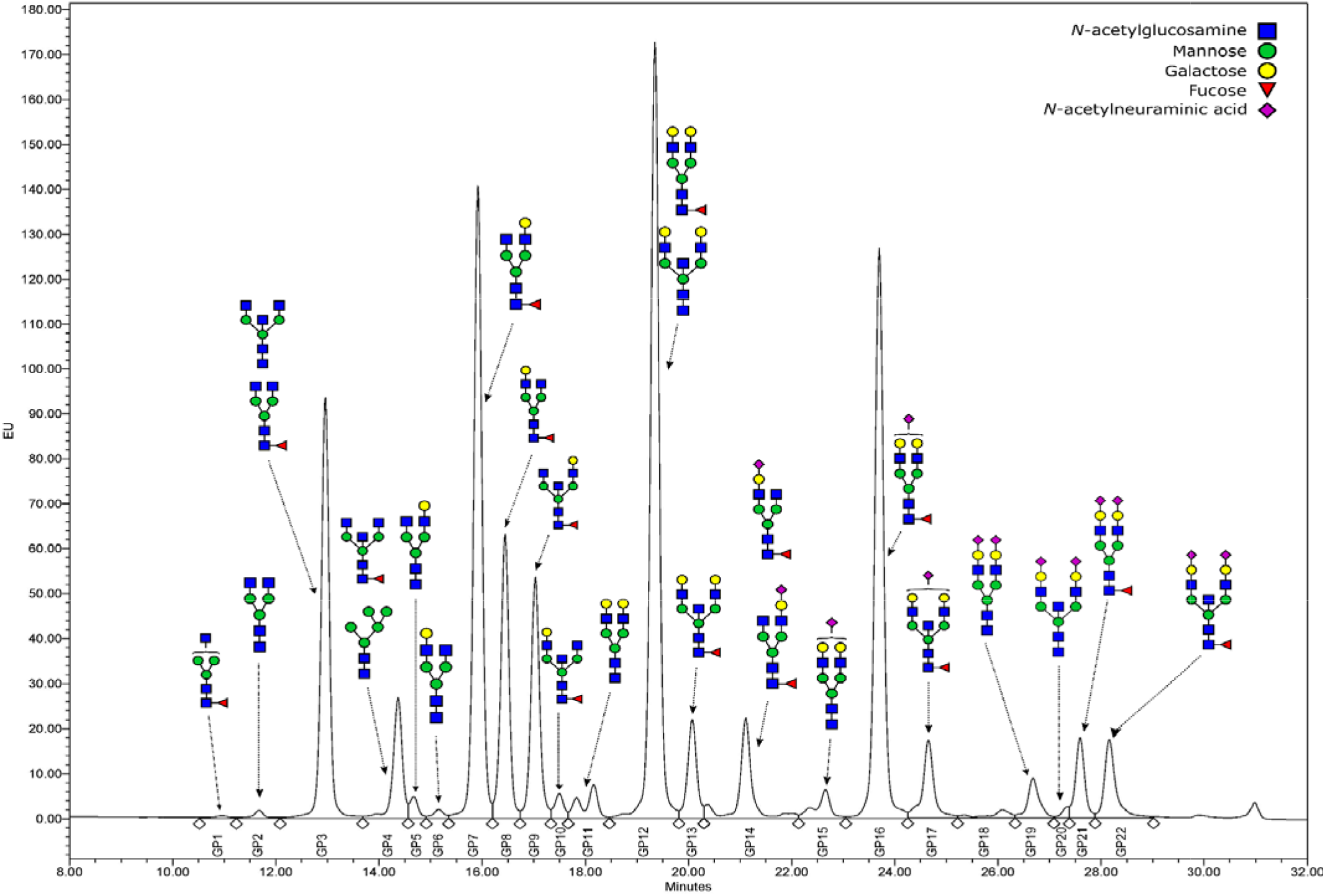
Representative HILIC-UPLC-FLR chromatographic profile of the immunoglobulin G RapiFluor-MS labelled N-glycome, with a graphic representation of the glycan structures corresponding to each glycan peak (GP). In case of multiple structures per GP, the upper structure is the major one, and the lower one is minor in abundance.

**Supplementary Figure 2.**
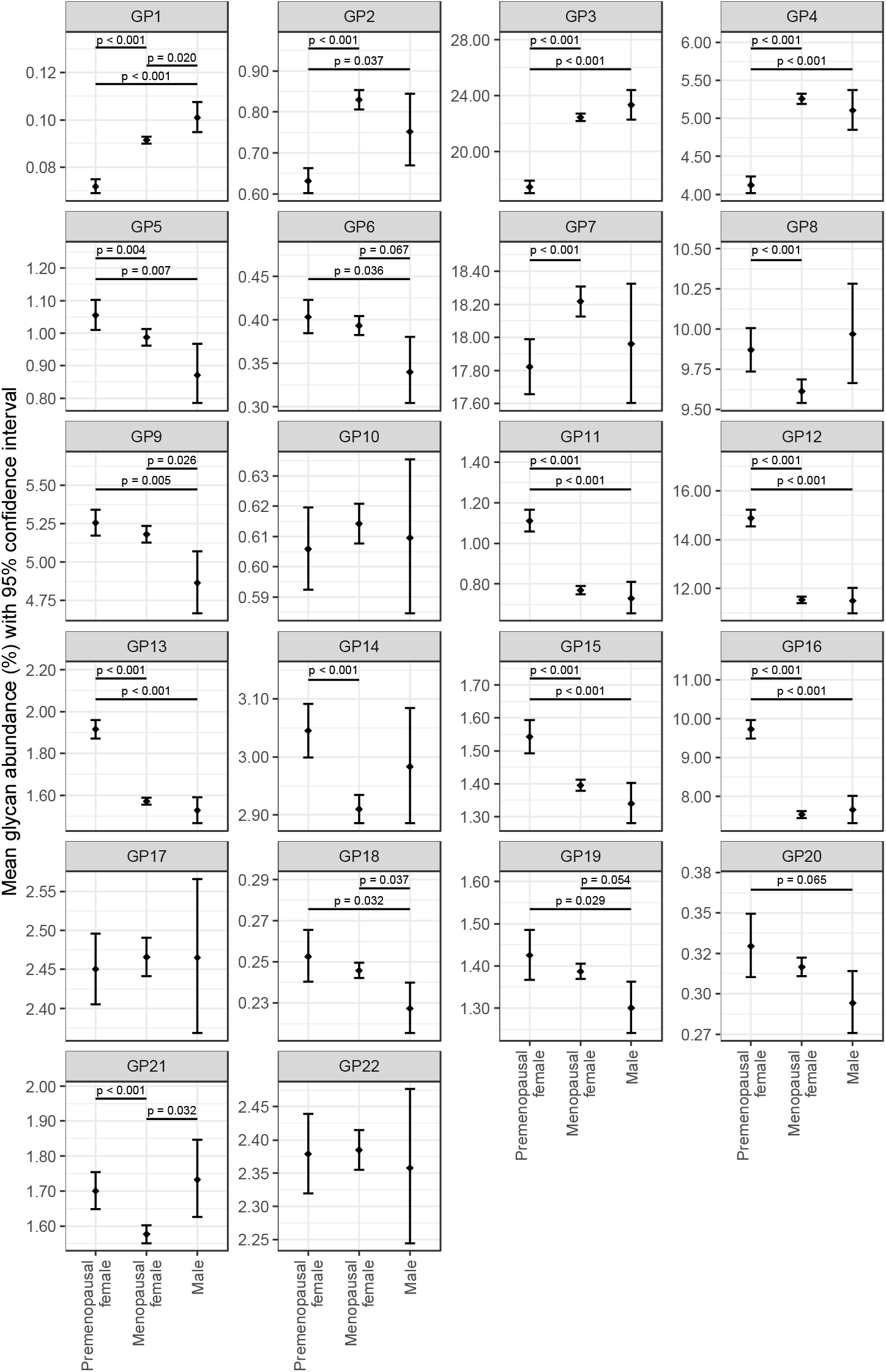
IgG glycome composition in premenopausal and menopausal women, and men. Mean glycan abundances (% of total IgG N-glycome) and corresponding 95% confidence intervals were estimated from fitted mixed model with logit transformed glycan as dependent variable and sex, menopausal status (nested within sex), age and age-menopausal status interaction as fixed factors, as well as family ID and individual ID (nested within family ID) as random intercepts and age as random slope. Only adjusted post-hoc p values less then 0.1 are shown.

**Supplementary Figure 3.**
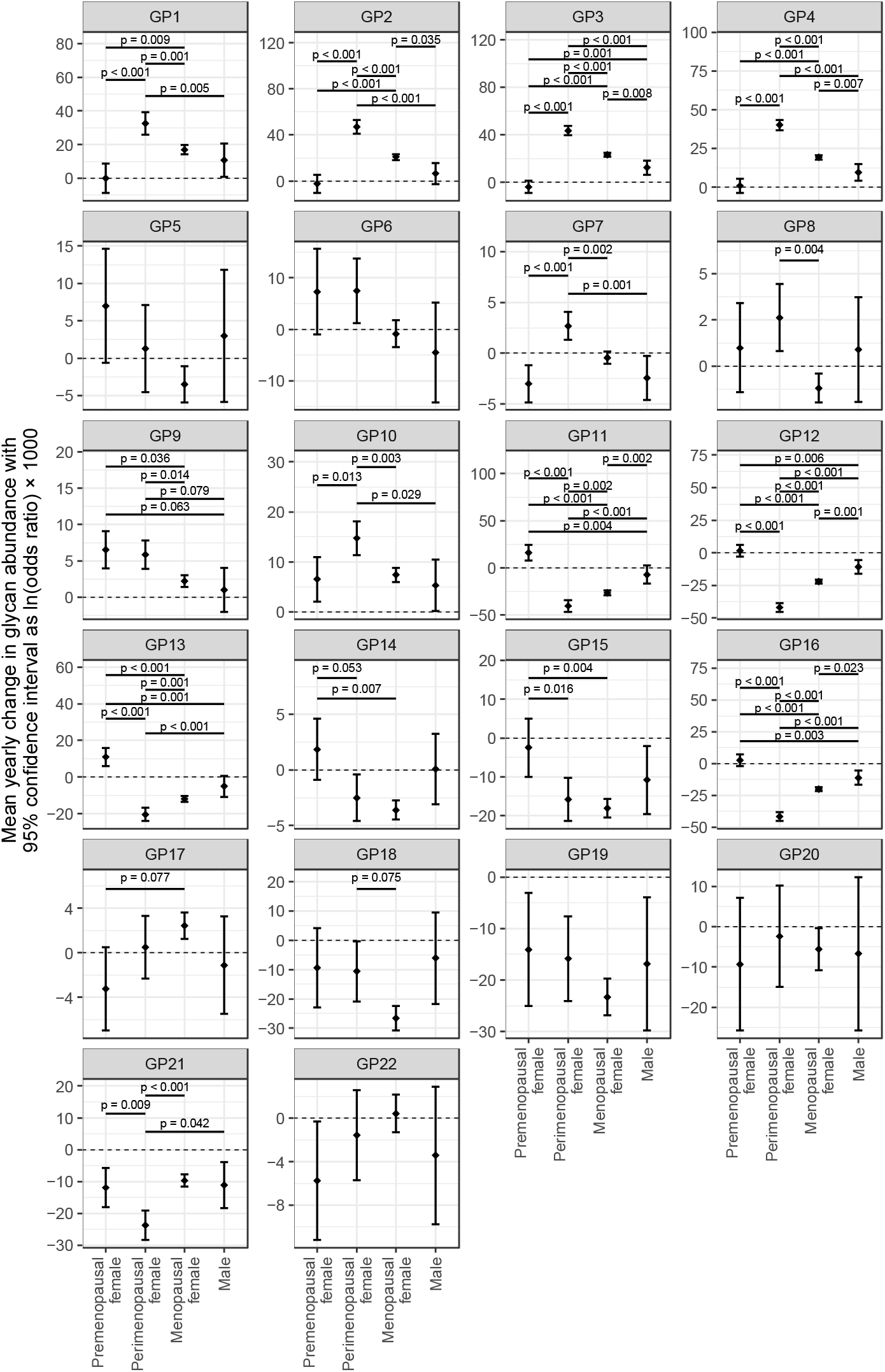
Average annual change of IgG glycans in females during the perimenopause period, females in pre- or post-menopause, and males. Mean yearly change in glycan abundances and corresponding 95% confidence intervals were estimated from fitted mixed model with logit transformed glycan as dependent variable and sex, menopausal status (nested within sex), age and age-menopausal status interaction as fixed factors, as well as family ID and individual ID (nested within family ID) as random intercepts and age as random slope. Only adjusted post-hoc p values less then 0.1 are shown.

